# Lack of genetic evidence for a role of SLC25A46 in alpha-synucleinopathies

**DOI:** 10.64898/2026.01.30.26344974

**Authors:** Han Yu, Sitki Cem Parlar, Konstantin Senkevich, Emma N. Somerville, Zhao Zhang, Lang Liu, Meron Teferra, Jamil Ahmad, Farnaz Asayesh, Guy A. Rouleau, Ziv Gan-Or

## Abstract

**Background:** The *SLC25A46* gene encodes a mitochondrial carrier protein previously implicated in neuropathy and optic atrophy. Biallelic variants in *SLC25A46* have been described in patients with Parkinson’s disease (PD) with optic atrophy, but the evidence supporting a role in PD remains limited.

**Objective:** To assess whether *SLC25A46* variants contribute to PD, REM sleep behavior disorder (RBD), or Dementia with Lewy Bodies (DLB).

**Methods:** We examined common variants using four representative PD genome-wide association studies (GWAS) and an RBD GWAS and applied Summary-data-based Mendelian Randomization (SMR) to evaluate whether genetically regulated expression of *SLC25A46* shows a causal association with the risk of PD or RBD. Rare variant analyses were conducted in four cohorts of European descent: Accelerated Medicines Partnership: Parkinson’s Disease (AMP-PD) PD (3,051 PD, 3,667 controls), UK Biobank (3,267 PD, 14,939 proxy, 54,800 controls), RBD (1,376 RBD, 2,580 controls), and AMP-PD DLB (2,605 DLB, 1,894 controls). Optimal Sequence Kernel Association test (SKAT-O) and meta-analysis were used to assess rare variants.

**Results:** No associations were observed between *SLC25A46* variants and PD, RBD, or DLB. SMR analyses revealed no evidence supporting a causal relationship between *SLC25A46* expression and PD or RBD risk. Rare variant burden analyses did not identify significant associations after multiple-testing correction across cohorts or meta-analyses.

**Conclusion:** *SLC25A46* variants showed no evidence of association, suggesting the gene does not play a major role in PD, RBD, or DLB risk.

## 1. Introduction

Parkinson’s Disease (PD) is a common neurodegenerative movement disorder, caused by a combination of genetic and environmental factors, with the normal process of aging (1, 2). Common variants associated with PD contribute 26 - 36% of its calculated heritability (3), yet rare genetic variants also contribute to PD. In particular, several autosomal recessive forms of PD have been well established, including those caused by mutations in *PRKN, PINK1* and *PARK7*, highlighting the contribution of rare gene defects to disease pathogenesis (4).

Previous studies have shown that mutations in *SLC25A46*, encoding a mitochondrial carrier protein, could cause peripheral neuropathy and optic atrophy (5). A possible link between recessive *SLC25A46* variants and PD with optic atrophy has been suggested (6). A recent study evaluated *SLC25A46* variation in idiopathic PD using the Global Parkinson’s Genetics Program (GP2) and Accelerated Medicines Partnership: Parkinson’s Disease (AMP-PD) whole-genome sequencing (WGS) data, supplemented by a large multi-ancestry genotype-imputed GP2 dataset, and found no strong evidence for an association with PD (7). However, because much of the analyses relied on imputed genotypes and sequencing-based gene-level burden evaluation was largely restricted to the AMP-PD PD cohort, the cumulative contribution of rare *SLC25A46* variants to PD risk remains to be further clarified.

In the present study, we aimed to evaluate the association of *SLC25A46* variants with PD, REM sleep behavior disorder (RBD), and Dementia with Lewy Bodies (DLB) across multiple cohorts.

## 2. Methods

### 2.1 Study Population

For common variants analysis, we used summary statistics from the two largest European PD GWASs (3, 8), the multi-ancestry and Asian PD GWASs (9, 10), and the RBD GWAS (11).

For summary-data–based Mendelian randomization (SMR) analyses, we used summary-level expression quantitative trait loci (eQTL) data. The exposure datasets included cortex cis-eQTL data from BrainMeta v.2 (12) (N = 2,443), and the Genotype-Tissue Expression (GTEx) v.8 (13) cis-eQTL data across caudate (N = 194), brain cortex (N = 205), substantia nigra (N = 114), and whole blood (N = 670). The outcome datasets were derived from the GWAS summary statistics described above. We used linkage disequilibrium (LD) reference data from the European-ancestry population of the 1000 Genomes Project Phase 3 (https://zenodo.org/records/10515792) to account for LD patterns among single-nucleotide polymorphisms (SNPs).

To analyze rare variants, we performed analyses in four cohorts with available WGS data: 1) a PD WGS cohort consisting of 3,051 cases and 3,667 controls obtained from the Accelerating Medicines Partnership® (AMP®) Parkinson’s Disease (AMP-PD) (https://amp-pd.org/) program, 2) UK biobank (UKBB) WGS cohort with 3,267 PD cases, 14,939 proxy cases, and 54,800 controls, which was accessed using Neurohub (https://www.mcgill.ca/hbhl/neurohub), 3) an RBD WGS cohort of 1,376 RBD cases and 2,580 controls collected through the International RBD Study Group (IRBDSG) from multiple centers across Europe and North America, sequenced at McGill University (11), and 4) a DLB WGS cohort consisting of 2,605 DLB cases and 1,894 controls obtained from the AMP-PD program. PD patients in the AMP-PD cohort and UKBB cohort were diagnosed by movement disorder specialists according to UK Brain Bank criteria (14) or MDS clinical diagnostic criteria (15). RBD was diagnosed with video-polysomnography according to the International Classification of Sleep Disorders (ICSD) version 2 or 3 (16, 17). DLB patients in the AMP-PD cohort were diagnosed by McKeith consensus criteria (18) or Emre consensus criteria (19).

Informed consent forms were signed by all participants before entering the studies and study protocols were approved by the institutional review boards.

### 2.2 Common Variant Association Analysis

To examine whether common variants in *SLC25A46* may be associated with PD or RBD, we created regional gene locus zoom plots using the online tool *LocusZoom* (20) for the *SLC25A46* loci with ±100 kb around the gene for each of the GWAS summary statistics.

### 2.3 Summary-data-based Mendelian Randomization

To investigate potential causal relationships between *SLC25A46* gene expression and disease risk, we conducted SMR analyses. Significant cis-eQTLs (p < 5 × 1e-8) from the BrainMeta and GTEx datasets were selected and restricted to variants within a ±1000kb cis-window around the gene. The heterogeneity in dependent instruments (HEIDI) test was applied to distinguish true causal effects from associations driven by LD.

### 2.4 Data Quality Control

Initial quality control procedures of AMP-PD cohorts were performed as described by AMP-PD (https://amp-pd.org/whole-genome-data) (21). Across all cohorts, additional QC was applied at both sample and variant levels.

At the genotype and variant level, low-quality genotypes were filtered based on genotype quality (GQ) and sequencing depth of coverage (DP), multi-allelic variants and variants with genotype missingness greater than 5% were excluded. Analyses were restricted to rare variants using a minor allele frequency (MAF) threshold of 0.01. Specifically, variants with DP < 25 were excluded in the two AMP-PD cohorts using bcftools (22). For the UKBB cohort, we used the Genome Analysis Toolkit (GATK, v4.2.5) (23) to filter out genotypes with GQ < 25 or DP < 25. For the RBD WGS data, variants were excluded for GQ < 20 or DP < 10, case control missingness (p < 1e-4), haplotype missingness (p < 1e-4), deviation from the Hardy Weinberg equilibrium in controls (p < 1e-4), using PLINK v1.9 (24).

At the sample level, analyses were restricted to individuals of European ancestry, and related individuals were excluded to retain unrelated samples. Samples with known quality issues (e.g., aneuploidy, genotype missingness > 5%, heterozygosity outliers, or discordance between genetic and reported sex) were removed, as applicable to each cohort.

Alignment of all cohorts was performed using the human reference genome (hg38). Using PLINK v1.9, we extracted the *SLC25A46* locus from each dataset based on its appropriate genomic coordinates (chr5:110,739,007-110,765,157).

### 2.5 Annotations and Statistical Analysis

To functionally annotate genetic variants in all cohorts, we used the Ensembl Variant Effect Predictor (VEP) (25). To assess the cumulative effect of multiple rare *SLC25A46* variants on the risk of PD, RBD, or DLB, we conducted gene-based burden analysis using optimized sequence kernel association test (SKAT-O) implemented in the SKAT R package (26). To explore whether variants with different functional characteristics contribute differently to association signals, variants were stratified into five categories: (1) All rare: all rare variants, (2) Non-synonymous: all non-synonymous variants, (3) Loss-of-function (LOF): all stop-gained, frameshift, start/stop lost, splice-site variants on splicing acceptor and donor sites, exon loss, and transcript ablation, (4) AlphaMissense pathogenic variants: missense variants scored by the AlphaMissense deep learning model and predicted to be likely pathogenic, and (5) Combined Annotation Dependent Depletion (CADD) ≥ 20: variants with CADD PHRED scores ≥ 20, representing the top 1% of potentially deleterious variants. For each variant category, we performed a PD meta-analysis between the AMP-PD PD cohort and the UKBB cohort using metaSKAT R package (27). Sex, age, and the top five principal components were included as covariates to account for potential confounding effects associated with these variables. To minimize the likelihood of false-positive findings, we applied false discovery rate (FDR) correction (28) to all P-values. The code supporting the findings of this study will be made publicly available on GitHub upon publication at: https://github.com/Hannah-Yu-0816/SLC25A46.

## 3. Results

LocusZoom visualization of common variants (MAF > 0.01) in *SLC25A46* using summary statistics from four representative PD GWASs and the RBD GWAS revealed no significant associations (p < 5 × 1e-8) with PD or RBD risk (Figure 1).

**Figure 1.**
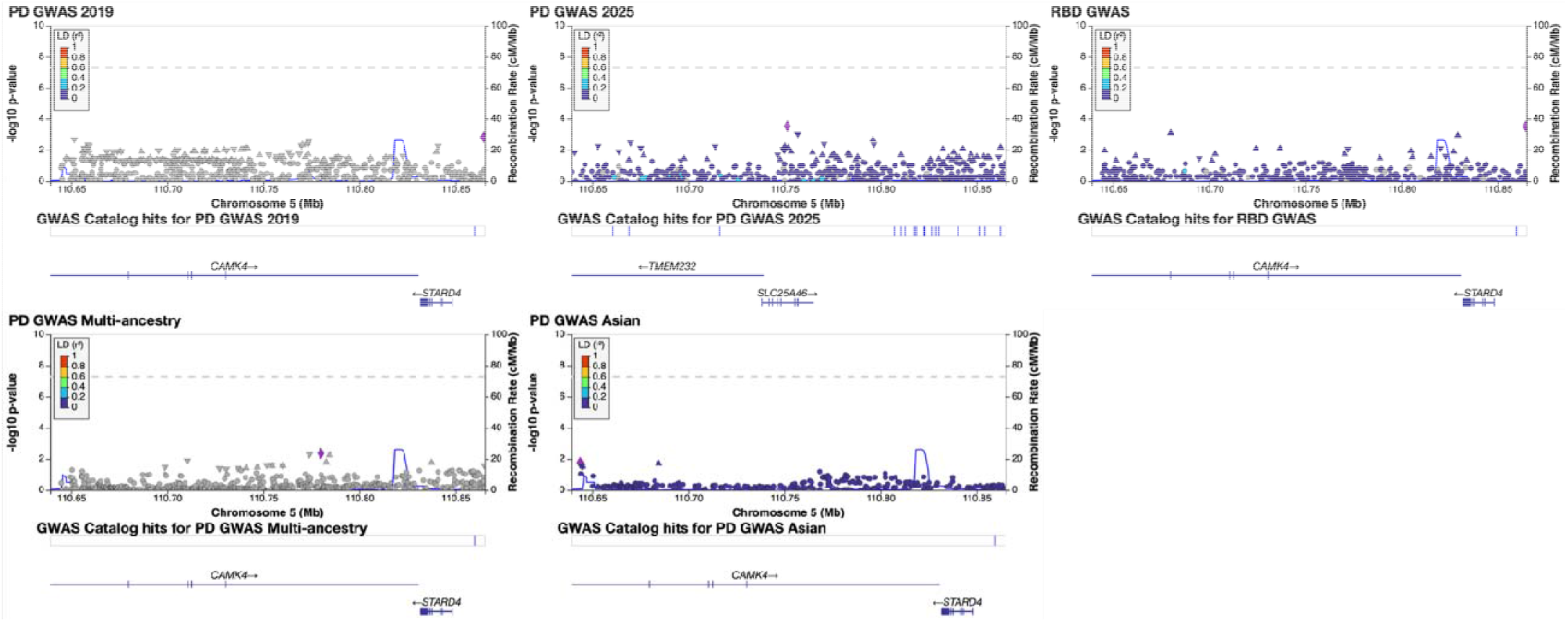
LocusZoom plots of *SLC25A46* gene (±100 kb) in the two largest European Parkinson’s disease (PD) genome-wide association study (GWAS), the multi-ancestry and Asian PD GWASs, and the REM sleep behavior disorder GWAS The lead SNP (i.e. the variant with the lowest P-value within each GWAS) is shown in purple. Variants are colored based on their linkage disequilibrium (LD) with the lead SNP, while grey variants indicate SNPs with no available LD information. The punctuated line represents the GWAS level of significance (P < 5 × 10e^-8^). X-axis: Chromosomal position (in Mb). Y-axis: Negative log10 of the P-values from GWAS.

Across all SMR analyses, we observed no statistically significant associations between *SLC25A46* expression and the risk of PD or RBD. None of the tested signals met the nominal significance threshold (p < 0.05) (Supplementary Table S1). The SMR analysis does not support a causal relationship between *SLC25A46* expression and susceptibility to PD or RBD.

The number of cases and controls after QC and the demographic information for each cohort can be found in Table 1. In the AMP-PD PD cohort, 887 rare variants were identified in the analysis, including 29 non-synonymous variants, 20 CADD ≤ 20 variants, one LOF variant, and 10 AlphaMissense likely pathogenic variants. In the UKBB cohort, 2094 rare variants were identified in the analysis, including 84 non-synonymous variants, 65 CADD ≤ 20 variants, 10 LOF variants, and 31 AlphaMissense likely pathogenic variants. In the RBD cohort, 677 rare variants were identified in the analysis, including 18 non-synonymous variants, 11 CADD ≤ 20 variants, one LOF variant, and five AlphaMissense likely pathogenic variants. In the AMP-PD DLB cohort, 759 rare variants were identified, including 20 non-synonymous variants, 12 CADD ≤ 20 variants, one LOF variant, and four AlphaMissense likely pathogenic variants (Table 2). The detailed information for the rare variants in each cohort can be found in Supplementary Tables S2-S5.

**Table 1.** Cohort demographics and individuals analyzed after quality control.

| Cohort | N Cases | N Controls | Status | Males (%) | Females (%) | Mean Age (±SD) |
| --- | --- | --- | --- | --- | --- | --- |
| RBD | 1376 | 2580 | Cases | 82.4 | 17.6 | 66.9 (9.2) |
|  |  |  | Controls | 50.7 | 49.3 | 69.9 (13.4) |
| AMP-PD<br>DLB | 2605 | 1894 | Cases | 63.5 | 36.5 | 77.0 (8.48) |
|  |  |  | Controls | 50.4 | 29.6 | 72.7 (13.4) |
| UKBB | 18206 | 54800 | Cases | 46.5 | 53.5 | 56.8 (8.00) |
|  |  |  | Controls | 46.6 | 53.4 | 59.2 (7.03) |
| AMP-PD<br>PD | 3051 | 3667 | Cases | 62.4 | 37.6 | 64.4 (9.74) |
|  |  |  | Controls | 46.7 | 53.3 | 67.9 (13.7) |

N, numbers; SD, standard deviation; RBD, REM sleep behavior disorder; DLB, Dementia with Lewy Bodies; UKBB, UK Biobank; AMP-PD, Accelerated Medicines Partnership: Parkinson's Disease

**Table 2.** SKAT-O analyses of rare variants (MAF < 0.01)

|  | RBD |  | AMP-PD<br>DLB |  | UKBB |  | AMP-PD PD |  | Meta-analysis of UKBB and AMP-PD PD |
| --- | --- | --- | --- | --- | --- | --- | --- | --- | --- |
| Variant category | N Variants | P-value | N Variants | P-value | N Variants | P-value | N Variants | P-value | P-value |
| All rare | 677 | 0.928 | 759 | **0.031** | 2094 | 0.591 | 887 | 0.687 | 0.717 |
| Non-synonymous | 18 | 0.275 | 20 | 0.581 | 84 | 0.488 | 29 | 0.600 | 0.497 |
| AlphaMissense likely pathogenic | 5 | 0.692 | 4 | 0.080 | 31 | 1 | 10 | 0.126 | 0.869 |
| Loss of function | 1 | 0.092 | 1 | 0.474 | 10 | 0.593 | 1 | 0.490 | 0.516 |
| CADD <sup>3</sup> 20 | 11 | 0.299 | 12 | 0.456 | 65 | 0.155 | 20 | 0.379 | 0.148 |

Note: P-value presented without FDR adjustment, as no P-values survived after correction.
SKAT-O, Optimal Sequence Kernel Association Test; MAF, minor allele frequency; RBD, REM sleep behavior disorder; DLB, Dementia with Lewy Bodies; UKBB, UK Biobank; AMP-PD, Accelerated Medicines Partnership: Parkinson's Disease; N, numbers; SD, standard deviation; CADD, Combined Annotation Dependent Depletion

No statistically significant associations were observed between rare variants and PD or RBD across any of the variant categories in any cohort or in the meta-analysis of PD. A nominal association of all rare variants in *SLC25A46* with DLB was found in the AMP-PD DLB cohort (P = 0.031) (Table 2), but it was not statistically significant after FDR correction.

We also investigated whether biallelic carriers of rare *SLC25A46* variants are overrepresented in PD. We identified two controls and one case who carried two rare CADD ≤ 20 and non-synonymous variants in the AMP-PD PD cohort, and four controls who carried two rare non-synonymous variants in the UKBB cohort (Supplementary Table S6). No PD cases with a diagnosis of optic atrophy were identified in the UKBB cohort.

## 4. Discussion

In the current study, we performed a comprehensive genetic association testing of both common and rare variants in *SLC25A46* to evaluate their potential contribution to the risk of PD, RBD, and DLB. Across all analyses, we did not observe any significant associations, suggesting that genetic variation in *SLC25A46* does not play a major role in the risk of either PD, RBD, or DLB in the studied cohorts. It is possible that biallelic *SLC25A46* variants lead to a specific subtype of PD and optic atrophy, yet it is also possible that they only cause optic atrophy, and the previously reported individuals with PD and optic atrophy had PD due to other reasons.

The biological rationale for investigating *SLC25A46* arises from its critical function in mitochondrial dynamics and neuronal maintenance, given that *SLC25A46* encodes an outer mitochondrial membrane carrier protein involved in regulating cristae structure and mitochondrial fission-fusion balance (29, 30). Subsequent studies further demonstrated that pathogenic mutations in this gene have been associated with optic atrophy, axonal neuropathy, and other mitochondrial disorders (31). Bitetto et al. described two individuals with biallelic *SLC25A46* mutations who presented Parkinsonism plus optic atrophy, suggesting potential phenotypic overlap between mitochondrial disorders and PD (6). Consistent with our findings, Schneider et al. reported no evidence for a major contribution of rare *SLC25A46* variants to idiopathic PD risk (7). However, that study focused on idiopathic PD and primarily evaluated single-variant signals and sequencing-based gene-level burden within the AMP-PD PD cohort. By extending the analysis to additional and larger cohorts, incorporating systematic rare-variant aggregation across functional categories, and examining RBD and DLB as a prodromal phenotype, our study provides complementary and independent evidence strengthening the overall genetic evidence base for *SLC25A46* in PD, RBD, or DLB. Taken together, our findings suggest that *SLC25A46* variation is unlikely to represent an important risk factor for PD, RBD, or DLB, even though *SLC25A46* dysfunction may cause rare mitochondrial syndromes featuring Parkinsonism and optic atrophy.

Our study has several limitations. First, although we included some of the largest PD, RBD, and DLB sequencing datasets to date, the statistical power to detect associations with very rare variants remains limited. Second, our study was restricted to individuals of European ancestry due to data availability, and potential population-specific effects cannot be excluded. Additional studies in other populations are required.

To conclude, our study does not provide evidence for involvement of *SLC25A46* variants in PD, RBD, or DLB. However, future work combining population genetics and functional studies will be valuable to elucidate how mitochondrial dynamics genes such as *SLC25A46* interact with broader pathways of neurodegeneration.

## 5. Acknowledgements

We would like to thank the research participants for contributing to this study. ZGO is supported by the Fonds de recherche du Québec–Santé (FRQS) Chercheurs-boursiers award and is a William Dawson Scholar. This research used the NeuroHub infrastructure and was undertaken thanks in part to funding from the Canada First Research Excellence Fund, awarded through the Healthy Brains, Healthy Lives initiative at McGill University, Calcul Québec and Compute Canada. This research has been conducted using the UK Biobank Resource under Application Number 45551. The UKBB cohort was accessed using Neurohub (https://www.mcgill.ca/hbhl/neurohub). Data used in the preparation of this article were obtained from the Accelerating Medicine Partnership® (AMP®) Parkinson’s Disease (AMP-PD) Knowledge Platform. For up-to-date information on the study, visit https://www.amp-pd.org. The AMP® PD program is a public-private partnership managed by the Foundation for the National Institutes of Health and funded by the National Institute of Neurological Disorders and Stroke (NINDS) in partnership with the Aligning Science Across Parkinson’s (ASAP) initiative; Celgene Corporation, a subsidiary of Bristol-Myers Squibb Company; GlaxoSmithKline plc (GSK); The Michael J. Fox Foundation for Parkinson’s Research; Pfizer Inc.; AbbVie Inc.; Sanofi US Services Inc.; and Verily Life Sciences. ACCELERATING MEDICINES PARTNERSHIP and AMP are registered service marks of the U.S. Department of Health and Human Services. Clinical data and biosamples used in preparation of this article were obtained from the (i) Michael J. Fox Foundation for Parkinson’s Research (MJFF) and National Institutes of Neurological Disorders and Stroke (NINDS) BioFIND study, (ii) Harvard Biomarkers Study (HBS) and the Stephen & Denise Adams Center for Parkinson’s Disease Research of Yale School of Medicine (CPDR-Y), (iii) National Institute on Aging (NIA) International Lewy Body Dementia Genetics Consortium Genome Sequencing in Lewy Body Dementia Case-control Cohort (LBD), (iv) MJFF LRRK2 Cohort Consortium (LCC), (v) NINDS Parkinson’s Disease Biomarkers Program (PDBP), (vi) MJFF Parkinson’s Progression Markers Initiative (PPMI), and (vii) NINDS Study of Isradipine as a Disease-modifying Agent in Subjects With Early Parkinson Disease, Phase 3 (STEADY-PD3) and (viii) the NINDS Study of Urate Elevation in Parkinson’s Disease, Phase 3 (SURE-PD3). BioFIND is sponsored by The Michael J. Fox Foundation for Parkinson’s Research (MJFF) with support from the National Institute for Neurological Disorders and Stroke (NINDS). The BioFIND Investigators have not participated in reviewing the data analysis or content of the manuscript. For up-to-date information on the study, visit michaeljfox.org/news/biofind. Genome sequence data for the Lewy body dementia case-control cohort were generated at the Intramural Research Program of the U.S. National Institutes of Health. The study was supported in part by the National Institute on Aging (program #: 1ZIAAG000935) and the National Institute of Neurological Disorders and Stroke (program #: 1ZIANS003154). The Harvard Biomarker Study (HBS) is a collaboration of HBS investigators [full list of HBS investigators found at https://www.bwhparkinsoncenter.org/biobank/ and funded through philanthropy and NIH and Non-NIH funding sources. The Stephen & Denise Adams Center for Parkinson’s Disease Research of Yale School of Medicine is funded through philanthropy and NIH and non-NIH funding sources. The HBS and CPDR-Y Investigators have not participated in reviewing the data analysis or content of the manuscript. Data used in preparation of this article were obtained from The Michael J. Fox Foundation sponsored LRRK2 Cohort Consortium (LCC). The LCC Investigators have not participated in reviewing the data analysis or content of the manuscript. For up-to-date information on the study, visit https://www.michaeljfox.org/biospecimens). PPMI is sponsored by The Michael J. Fox Foundation for Parkinson’s Research and supported by a consortium of scientific partners: [list the full names of all of the PPMI funding partners found at https://www.ppmi-info.org/about-ppmi/who-we-are/study-sponsors]. The PPMI investigators have not participated in reviewing the data analysis or content of the manuscript. For up-to-date information on the study, visit www.ppmi-info.org. The Parkinson’s Disease Biomarker Program (PDBP) consortium is supported by the National Institute of Neurological Disorders and Stroke (NINDS) at the National Institutes of Health. A full list of PDBP investigators can be found at https://pdbp.ninds.nih.gov/policy. The PDBP investigators have not participated in reviewing the data analysis or content of the manuscript. The Study of Isradipine as a Disease-modifying Agent in Subjects With Early Parkinson Disease, Phase 3 (STEADY-PD3) is funded by the National Institute of Neurological Disorders and Stroke (NINDS) at the National Institutes of Health with support from The Michael J. Fox Foundation and the Parkinson Study Group. For additional study information, visit https://clinicaltrials.gov/ct2/show/study/NCT02168842. The STEADY-PD3 investigators have not participated in reviewing the data analysis or content of the manuscript. The Study of Urate Elevation in Parkinson’s Disease, Phase 3 (SURE-PD3) is funded by the National Institute of Neurological Disorders and Stroke (NINDS) at the National Institutes of Health with support from The Michael J. Fox Foundation and the Parkinson Study Group. For additional study information, visit https://clinicaltrials.gov/ct2/show/NCT02642393. The SURE-PD3 investigators have not participated in reviewing the data analysis or content of the manuscript. Additionally, the G-Can (GBA1-Canada) Initiative, an open-science collaborative initiative aimed at addressing GBA1-associated neurodegeneration, has made contributions to this research.

## 6. Author contribution

HY conducted the analysis, interpreted the results, and wrote the manuscript. SCP and KS assisted with computational analysis and support. ENS, ZZ, and LL contributed to the WGS data quality control process. ZG-O and KS supervised the study and contributed to the interpretation of the results. All authors reviewed, approved and contributed to editing the final manuscript.

## 7. Statements and declarations

### 7.1 Ethical considerations

Ethics approval for the research study was granted by the McGill University Research Ethics Board.

### 7.2 Consent to participate

Informed consent forms were signed by all participants before entering the studies.

### 7.3 Consent for publication

Not applicable.

### 7.6 Data availability

PD risk GWAS summary statistics (Nalls et al. 2019) can be downloaded from the GWAS Catalog (https://www.ebi.ac.uk/gwas/, study accession GCST009325). PD risk GWAS summary statistics (GP2 2025) are publicly available at https://ndkp.hugeamp.org/research.html?pageid=a2f_downloads_280. Multi-ancestry PD risk GWAS summary statistics (Kim et al. 2024) are accessible via the Neurodegenerative Disease Knowledge Portal at https://ndkp.hugeamp.org/. Asian PD risk GWAS summary statistics (Foo et al. 2020) are available from the original study authors upon reasonable request. The iRBD summary statistics can be found in the GWAS Catalog (https://www.ebi.ac.uk/gwas/, study accession GCST90204200). eQTL data from BrainMeta version 2 cis-eQTL and Genotype-Tissue Expression (GTEx) version 8 cis-eQTL summary statistics are available at https://yanglab.westlake.edu.cn/software/smr/#DataResource. GWAS summary statistics for PD progression, the Montreal Cognitive Assessment (MoCA), and UPDRS Part 3 traits are publicly available at https://pdgenetics.shinyapps.io/pdprogmetagwasbrowser/. The data used in the preparation of this article were obtained from the AMP-PD Knowledge Platform (https://www.amp-pd.org) and the UKBB via Neurohub (https://www.mcgill.ca/hbhl/neurohub).

